# Plasma proteomic profiles of UK Biobank participants with Multiple Sclerosis

**DOI:** 10.1101/2023.07.25.23293146

**Authors:** Benjamin M Jacobs, Nicola Vickaryous, Gavin Giovannoni, Petroula Proitsi, Sheena Waters, Ruth Dobson

## Abstract

**Objective:** We aimed to describe plasma protein biomarkers of Multiple Sclerosis risk and to explore protein biomarkers of disease severity using radiological outcome measures.

**Methods:** Multiple Sclerosis cases and controls were identified in UK Biobank, a longitudinal cohort study of ∼500,000 British adults. Plasma proteins were assayed in ∼50,000 UK Biobank participants using the Olink proximity extension assay. We performed case-control association testing to examine the association between 2911 proteins and Multiple Sclerosis, using linear models adjusted for confounding covariates. Associations with radiological lesion burden and brain volume were determined in a subset of the cohort with available magnetic resonance imaging, using normalised T2-hyperintensity volume or whole brain volume as the outcome measure.

**Results:** 407 prevalent Multiple Sclerosis cases and 39,979 healthy controls were included. We discovered 72 proteins associated with Multiple Sclerosis at a Bonferroni-adjusted p-value of 0.05, including established markers such as Neurofilament Light Chain and Glial Fibrillary Acidic Protein. We observed a decrease in plasma Granzyme A, a marker of T cell and NK cell degranulation, which was specific to Multiple Sclerosis. Higher levels of plasma proteins involved in coagulation were associated with lower T2 lesion burden and preserved brain volume.

**Interpretation:** We report the largest plasma proteomic screen of Multiple Sclerosis, replicating important known associations and suggesting novel markers, such as the reduction in granzyme A. While these findings require external validation, they demonstrate the power of biobank-scale datasets for discovering new biomarkers for Multiple Sclerosis.

## Introduction

Blood-based protein biomarkers have several properties which make them attractive adjuncts in the diagnosis, monitoring, and prognostication of Multiple Sclerosis (MS). Proteins are often abundant in serum/plasma, stable in solution, and are straightforward to quantify. Furthermore plasma is easier to obtain than cerebrospinal fluid or Magnetic resonance imaging (MRI) scans, particularly if regular measurements are required for ongoing monitoring.

Plasma proteins do not currently feature routinely in the investigation or management of MS^1^, although plasma neurofilament light chain (NFL) is increasingly entering research and clinical practice. Prior studies have shown that plasma markers of neuroaxonal damage - NFL and glial fibrillary acidic protein (GFAP) - are elevated early in the MS disease course and show potential use for tracking disease activity and response to treatment^2,3^. However, neither of these markers is specific to MS^4–6^. Incorporating information from across the entire spectrum of plasma proteins - the proteome - may help to shed light on aspects of MS biology beyond neuronal loss, refine these tools for use on an individual basis, and suggest new targets for therapeutic intervention.

Previous efforts to explore the MS proteome have discovered proteins which correlate with either disease status or disease progression. However, the small sample size of these studies, differing technologies, and different protein targets have made it challenging to integrate these datasets^7–11^. A recent Swedish study compared the plasma proteome of MS cases and healthy controls using the Olink proximity extension assay applied to separate discovery (N_MS_=92, N_Control_=23) and replication (N_MS_=51, N_Control_=20) cohorts^12^, and found no significant differences after correction for multiple testing. An earlier study using a smaller set of proteins focussed on inflammation compared 111 MS cases and 46 healthy controls in the discovery cohort and a replication dataset of 94 cases and 47 controls. In the discovery cohort, oncostatin M (OSM) and hepatocyte growth factor (HGF) were increased in MS, whereas fibroblast growth factor 21 (FGF-21), fibroblast growth factor 23 (FGF-23), and cystatin-D (CST5 / CST-D) were decreased^9^. Of these signals, only the OSM and HGF signals replicated^9^. Another studycompared 90 MS cases and 20 controls in the Dutch GeneMSA cohort using the SomaLogic assay: the clearest difference between MS and controls was the downregulation of matrix metalloproteinase-3 (MMP3) in MS cases^7^. Studies of plasma proteomic correlates of disease severity have shown consistent associations with elevated markers of neuronal damage (GFAP and NFL) and myelin oligodendrocyte glycoprotein (MOG)^2,3,13^, but other markers have shown variable results between studies.

Recent advances have made it feasible to assay over a thousand different proteins in a sample simultaneously using either aptamer- or antibody-based technology. UK Biobank - a large long-term biobank study of >500,000 British adults - has recently applied the Olink proximity extension assay technology to measure the plasma level of ∼3000 proteins in >50,000 of its participants, providing an invaluable resource for examining the associations between genetics, protein levels, and health outcomes^14–16^. The prevalence of MS in UK Biobank is similar to the UK population (0.5%), and so this resource is also of significant value for MS proteomics research^17^.

In this study, we analysed proteomic data from MS cases and healthy controls in UK Biobank to search for proteomic signatures associated with MS disease status and with MS severity.

## Materials and methods

### Cohort

UK Biobank is a longitudinal cohort study of ∼500,000 British adults. Participants aged 40-69 were recruited between 2006 and 2010. A range of data is available for UKB participants, including self-reported lifestyle questionnaire data, linked electronic healthcare records, and genotyping/genome sequencing, described in detail elsewhere^18^. Plasma samples for proteomics were obtained at enrolment during the baseline visit. Plasma proteomics data covering a subset of the cohort were generated by the UK Biobank Pharma Proteomics Project^19,20^

### Case-control definitions

MS disease status was derived from the UK Biobank ‘first occurrence’ fields (UKB category 1712), which combine Hospital Episode Statistics (HES), Primary Care data, mortality records, and self-reported data to define the earliest report of each ICD10 code per participant in the dataset. MS cases were defined as participants with >=1 occurrence of the ICD10 code G35, an MS diagnostic code in primary care, HES, or self-reported MS. We distinguished prevalent and incident cases based on whether the first diagnostic code occurred before or within 10 years of recruitment to allow for diagnostic and recording lag. Using the same approach, we identified participants with neurodegenerative disorders - Parkinson’s Disease (G20), Alzheimer’s Disease (G30), Motor Neuron Disease (G12) - and other autoimmune disorders - Systemic Lupus Erythematosus (M32), Ankylosing Spondylitis (M45), Coeliac Disease (K90), Crohn’s Disease (K50), Ulcerative Colitis (K51), Rheumatoid Arthritis (M05), and psoriasis (L40).

We excluded cases with missing values for the date of earliest diagnostic code report or with age at diagnosis <= 10 years old. For the primary analysis, we included all prevalent MS cases and excluded controls with prevalent or incident diagnoses of any listed autoimmune / neurodegenerative conditions. Counts for each disease are shown in supplementary figure 1.

We excluded samples processed as part of the COVID or pilot batches to minimise biases due to the non-random sampling of the cohort^15^. These samples include controls selected for follow-up as part of ongoing COVID studies and so do not represent a random cohort selection. In addition, the risk of batch effects is higher with the pilot batch as there are a lower number of samples included, so these samples were also excluded, in line with previous analysis strategies employed by the UK Biobank Pharma Proteomics Project.

### Proteomics data

In April 2023, UK Biobank released proteomic assay data covering ∼55,000 participants^15^. The methods used to generate these data and in-house quality control have been previously described^15^. In brief, UKB performed multiplexed proteomic assays on ∼55,000 plasma samples obtained (for the majority) at baseline visits using the Olink platform, which exploits dual barcoded antibody technology to produce specific and semiquantitative readouts for 2923 protein assays in multiplex, selected to provide coverage across a range of systems and diseases. We used proteomic data derived from blood taken at the baseline visit. From the 2923 protein assays available, we excluded proteins with >20% of values missing (n=12), resulting in 2911 protein assays retained for analysis. Missing values in the remaining 2911 proteins were imputed with the mean value for the whole cohort.

### Statistical analysis

#### 1. Case-control comparison

To assess the relationship between plasma protein levels and Multiple Sclerosis, we filtered the dataset to prevalent MS cases and healthy controls and constructed linear regression models for each protein of the form: Protein level (NPX; normalised protein expression) ∼ age + sex + age^2^ + age x sex + age^2^ x sex + batch + disease status (MS vs control)

The outcome for each regression model was the NPX value, a normalised value corresponding to the log_2_ fold change in protein expression between samples, i.e. a 1-point NPX difference equates to a ∼2x higher concentration of protein. Linear models were fitted using the *limma* R package, which uses empirical Bayes estimators to calibrate the per-protein variance using information from all proteins. We reported results as statistically significant if they surpassed a Bonferroni-adjusted alpha threshold of 0.05 (P < 1.7 x 10^-5^ = 0.05 / 2911).

We performed the following sensitivity analyses to explore the impact of various possible confounders:

- Adjustment for BMI: Protein level (NPX; normalised protein expression) ∼ age + sex + age^2^ + age x sex + batch + age^2^ x sex + BMI + disease status (MS vs control)
- Simplified model: Protein level (NPX; normalised protein expression) ∼ age + sex + batch + disease status (MS vs control)
- Matched case-control analysis: we repeated the analysis in a matched case-control subsection of the cohort, matching each MS case to four healthy controls on age at recruitment (rounded to the nearest year) and sex. We used the same model specification as for the primary analysis.

Although visual inspection of the top hits revealed normally-distributed protein levels, we performed a further sensitivity analysis in which protein levels were rank-inverse normalised prior to model fitting to ensure that regression assumptions were satisfied.

#### 2. MS-specific biomarkers

To determine whether these biomarkers were specific to MS, we then performed case-control association testing between each protein biomarker and a variety of neurodegenerative and autoimmune disorders. For these models, we used the same model specification as in the primary analysis (adjusting for age, sex, batch, age^2^, age x sex, and age^2^ x sex). For each disease, we again compared prevalent cases with healthy controls (without neurodegenerative or autoimmune disorders) and excluded incident cases.

We then considered whether the MS-associated proteins identified were MS-specific using a stringent definition:

- The protein must be associated with MS at *P_Bonferroni_*<0.05 and

- The protein must not show association in the same direction with **any** other tested autoimmune or neurodegenerative disease at a nominal *P*<0.1.

#### 3. Association with MS disease-modifying therapy

We defined medication usage using the UK Biobank field field 20003, which records medications reported by the participant at their baseline visit. We identified MS disease-modifying therapies from the list of medications, and considered medications prescribed to >=10 individuals in the proteomics dataset. The only MS DMT with >=10 treated patients was interferon – we combined various formulations (including avonex, rebif, and betaferon) into a single binary variable (treated vs untreated). We tested for associations with interferon use within the MS cohort by fitting models with *limma* of the form: Protein level (NPX; normalised protein expression) ∼ age + sex + age^2^ + age x sex + age^2^ x sex + batch + treatment status (interferon vs no interferon)

#### 4. Proteomic correlates of T2 lesion load and brain atrophy

To identify plasma protein biomarkers of lesion volume in MS, we used MRI scans acquired on a subset of ∼45,000 UK Biobank participants^21,22^. We used the pre-computed total volume of brain T2 hyperintensities (UK Biobank field ID 25781) derived from T2-weighted FLAIR images. To account for variation due to head size, we calculated the normalised T2 lesion volume by dividing total T2 lesion volume by the total intracranial volume (the sum of the brain volume and the CSF volume). Normalised T2 lesion volumes were rank-inverse normalised before model fitting to ensure linear model assumptions were satisfied. We then tested for association between the 2911 proteins and normalised T2 lesion volume within the prevalent MS cases. Using the same approach, we examined the proteomic correlates of brain atrophy in MS by assessing the association between each proteins and the total volume of grey and white matter normalised for head size (UK Biobank field ID 25009), a cross-sectional readout of brain volume. As MRI scans were captured after the baseline visit, for both of these analyses we included the age at scan as an additional covariate in the models in addition to age at MS diagnostic report (i.e. we adjusted for age at recruitment, sex, age at recruitment^2^, age at recruitment x sex, age at recruitment^2^ x sex, age at MS diagnostic code report, BMI at the time of scan, batch, and age at scan). We adjusted for the BMI at the time of scan for these models, rather than the BMI at baseline visit. We confirmed there was no correlation between age at recruitment and age at scan (Pearson’s correlation coefficient -0.04, *P*=0.6) prior to model fitting to quantify the risk of multicollinearity. We performed sensitivity analysis using simplified models, adjusting for age at scan, sex, and proteomics batch.

#### 5. Gene/protein set enrichment analysis

We performed gene/protein set enrichment analysis to infer which pathways were altered in MS versus healthy controls. To do so, we used the fast gene set enrichment analysis R package (fgseaR)^23^, which estimates empirical P values for enrichment using a permutation-based approach. We explored pathways for which there was a minimum of 30 proteins overlapping between our set of tested proteins and the gene/protein set, used 10,000 permutations, and stipulated a maximum gene/protein set size of 1000. Proteins were ranked using the product of the sign of the correlation coefficient and the negative log10 of the P value, i.e. the proteins with the smallest P values and positive beta coefficients were ranked top, and those with the smallest P values and negative beta coefficients were ranked bottom. We report statistically significant results at a False Discovery Rate of 5%. As a reference, we used pathways from the Kyoto Encyclopaedia of Genes and Genomes (KEGG)^24^.

### Replication and power calculations

To determine whether the MS-associated proteins we identified replicated in an external dataset, we downloaded the list of proteins assessed in Akesson *et al* 2023^12^, which compared MS and controls from two Swedish cohorts, a discovery cohort from Linköping University Hospital and a replication cohort from Karolinska University Hospital. After filtering to the set of overlapping proteins, we considered effects ‘concordant’ if the effect direction was the same, i.e. the protein was either up– or down-regulated in all three datasets (i.e. UK Biobank and these two Swedish cohorts).

Power calculations were performed by simulating a normally-distributed protein level with a standard deviation of 1 and a mean value of 1 in controls. We explored a range of true log-fold change values and set the mean in cases to this value, i.e. for a log-fold change of 0.2, we set the mean in cases to 2^0.2^ ∼ 1.15. We performed 1,000 bootstrap simulations over a range of plausible log-fold change values, also varying the number of cases and controls. For each simulation, we used a univariable linear regression model, regressing the normalised protein level on case/control status. Power at the 5% alpha level was defined as the proportion of bootstrap iteration with a linear regression P value of <0.05.

### Computing & code availability

All analyses were conducted in R version >4.2.2. This research utilised Queen Mary’s Apocrita HPC facility, supported by QMUL Research-IT^25^. All code used for analysis is available at https://benjacobs123456.github.io/ukb_proteomics/.

### Data availability

UK Biobank data are available on request from https://www.ukbiobank.ac.uk/. This research was conducted under approved application 78867.

### Reporting guidelines

This research was conducted in accordance with the STROBE guidelines on observational studies. The completed STROBE checklist is provided in the supplementary materials.

## Results

### Demographics

From the 52,701 participants with proteomic data derived from their baseline sample, we excluded 7,588 participants included as part of the COVID substudy or in the pilot batch, leaving 45,113 participants for analysis (supplementary figure 2). We selected healthy controls by excluding 1375 controls with incident or prevalent neurodegenerative disorders (PD, AD, or ALS), and 3239 controls with incident or prevalent other autoimmune diseases (Coeliac disease, Ankylosing Spondylitis, Rheumatoid Arthritis, Ulcerative Colitis, Crohn’s disease, Psoriasis). We excluded participants with a diagnosis of MS coded more than ten years after enrolment (i.e. ‘incident’ cases, n=29), leaving 407 prevalent MS cases and 39,979 healthy controls. Demographics of the included cases and controls are shown in table 1. The median age at MS report was 44.4 years old (IQR 16.8), broadly consistent with previous estimates of the peak incidence of MS derived from electronic health records^26^. Most MS cases (326, 80.0%) had at least two separate sources of diagnostic code report (i.e. two of self-report, primary care, or hospital episode statistics). Compared with controls the MS cohort was younger, more affluent, with a lower Body Mass Index (BMI), higher socio-economic status (the Townsend deprivation score was used as a proxy) with a higher proportion of women and self-reported White British individuals (Table 1).

### Plasma protein alterations in Multiple Sclerosis

To search for protein biomarkers of Multiple Sclerosis, we compared the plasma proteomic profiles of 407 prevalent MS cases and 39,979 healthy control participants in UK Biobank. Of the 2911 proteins tested, 72 were associated with Multiple Sclerosis at a stringent family-wise error rate of 5% (lJ_Bonferroni_<0.05, P<1.7x10^-5^). Thirteen were increased in MS, and 59 decreased in MS (figure 1A, supplementary table 1). Plasma samples from MS cases tended to have higher levels of the neuroaxonal damage marker Neurofilament light chain (NFL), chromogranin A (CHGA), Lysosomal Associated Membrane Protein 3 (LAMP3), interleukin-17 receptor beta (IL17RB), interleukin 22 (IL22), insulin-like growth factor binding protein 2 (IGFBP2), leukocyte immunoglobulin like receptor 4 (LILRB4), creatine kinase B (CKB), glial fibrillary acidic protein (GFAP), cathepsin F (CTSF), interleukin-15 (IL15), MER proto-oncogene tyrosine kinase (MERTK), and folate receptor alpha (FOLR1) than controls (table 2, supplementary table 1). Most of these associations (49/72, 68.1%) persisted after accounting for the impact of BMI on the proteome ^15^ – of the thirteen proteins elevated in the MS samples, three of these associations (FOLR1, CKB and IGFBP2) dissipated on adjustment for BMI. To account for possible confounding due to differences in age and sex, we repeated the analysis in a nested matched case-control cohort (407 MS and 1628 age and sex-matched controls). Despite the lower statistical power due to the smaller number of controls, we observed strong evidence for elevation of all ten MS-associated proteins (supplementary table 1). Sensitivity analyses adjusting for just age, sex, and batch, and transforming protein levels yielded similar results (supplementary table 1).

**Figure 1:**
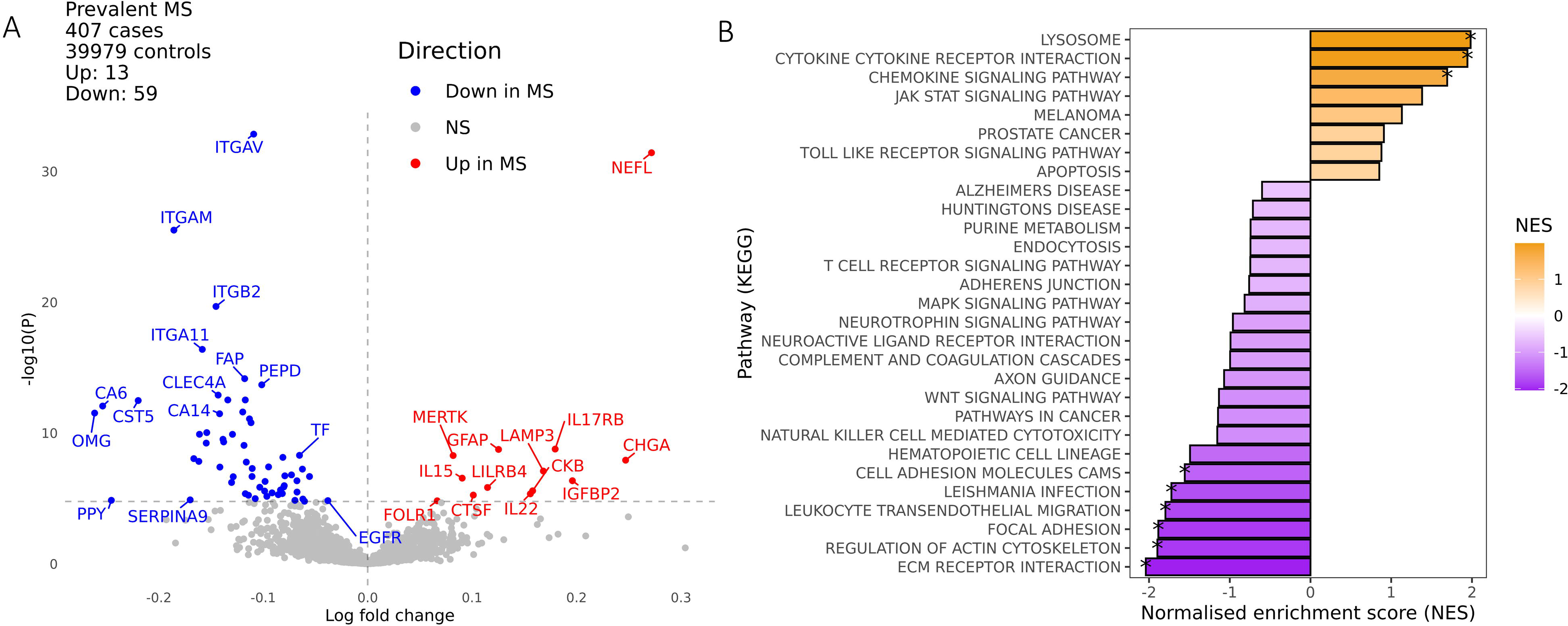
plasma proteomic analysis of Multiple Sclerosis. A - volcano plot displaying differences in plasma levels of proteins measured on the Olink proteomics assay between UK Biobank participants with (n=407) and without (n=39979) MS at the time of sampling. The x-axis indicates the log-fold change of the protein, with values above 0 indicating proteins present at higher levels in the MS cohort and those with values below 0 present at lower levels in the control cohort. The y-axis indicates the negative log of the P value for each protein, with higher values indicating a more statistically significant result. Proteins surpassing a Bonferroni-corrected threshold of 5% are shown in colour; other results are shown in grey. B - gene set enrichment analysis (GSEA) results summarising the pathway-level differences in the plasma proteome between MS and healthy controls. The x-axis shows the normalised enrichment score, with positive values equivalent to an increase in the pathway activity in MS and negative values suggesting decreased levels of pathway activity in MS. The y-axis shows the KEGG pathways tested. The asterisks indicate statistically significant results at an FDR of 5%.

Gene set enrichment analysis reaffirmed the enrichment of cytokines and cytokine receptors and lysosomal processing proteins in MS and the MS-associated decrease in proteins involved in leukocyte migration, interaction with the extracellular matrix, regulation of the actin cytoskeleton, and cell-cell adhesion (KEGG terms with normalised enrichment scores [NES] at FDR < 5%, figure 1B). We also observed suggestive evidence for an increase in proteins involved in JAK-STAT signalling (unadjusted *P* < 0.05).

### Replication of plasma proteomic signature

To validate these findings, we compared our results with recently published proteomics data from two MS cohorts assayed using the Olink platform^12^ (discovery dataset N_MS_=92, N_Control_=23, replication dataset N_MS_=51, N_Control_=20). Of the 49 MS-associated proteins we report which were robust to adjustment for BMI, 34 were assessed in these two cohorts (figure 2A & 2B). We observed directionally-concordant associations with MS in both the discovery and replication cohorts for 28/34 (82.4%) proteins, including most of the proteins increased in MS in UKB (IL17RB, NEFL, CTSF, MERTK, GFAP, LILRB4, and LAMP3), but not IL15. The only protein achieving more stringent statistical evidence for replication in both these datasets (i.e. nominal P value < 0.05) was Delta/Notch Like EGF Repeat Containing (DNER), which was decreased in all cohorts. Empirical power calculations (figure 2C) suggest that the power to observe an effect of a similar magnitude to the elevation in NFL (∼ log fold change of 0.2) was small in this study (10% for both the discovery and replication datasets at an alpha of 0.05). Therefore the directional concordance we observe can be interpreted as reasonable evidence for replication given the sample size.

**Figure 2:**
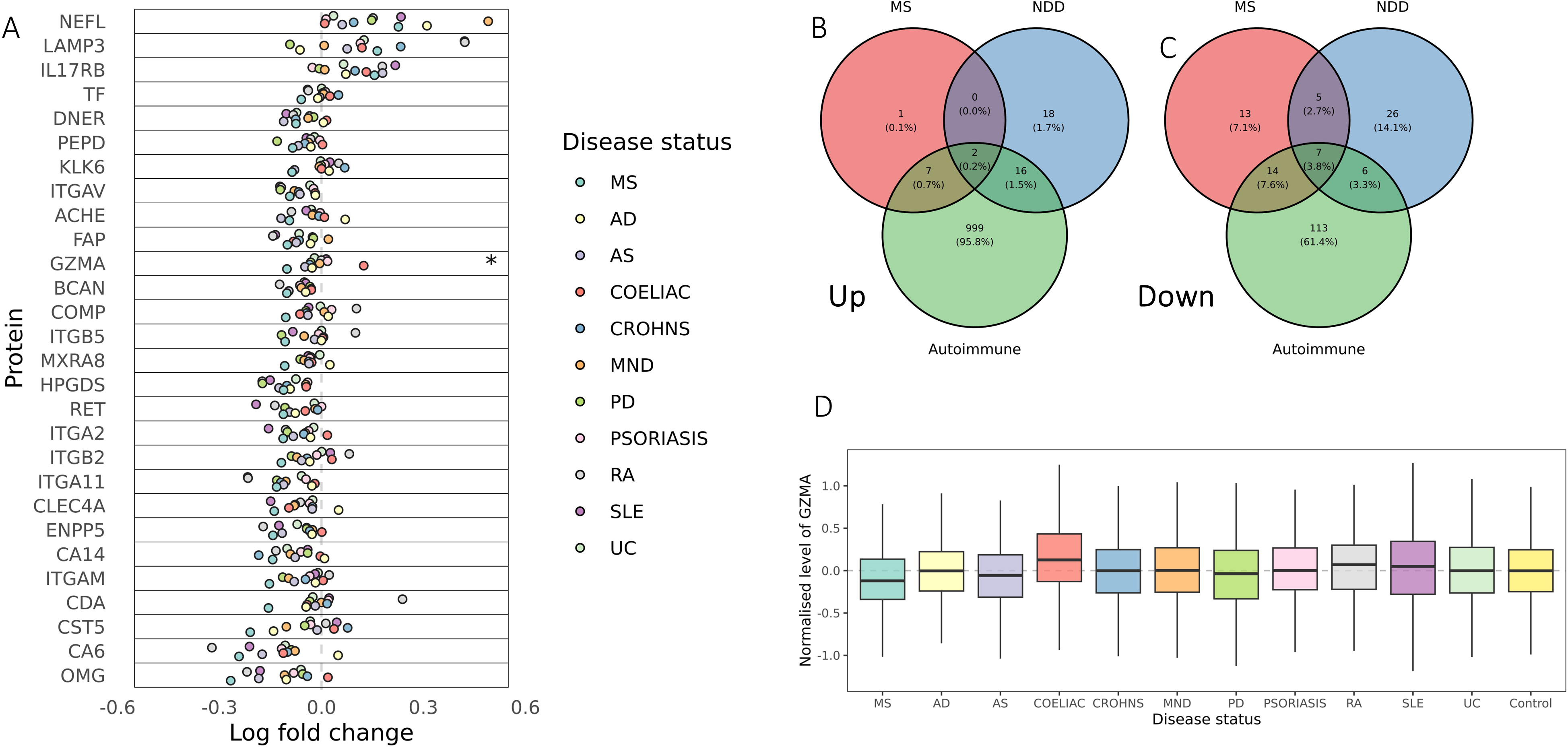
**external validation of plasma protein alterations in MS**. A: Beta-beta plots showing the log-fold change for each of the proteins identified as differentially-expressed in MS plasma in UKB & in a recent study of similar design using the Olink platform. The x axis shows the log-fold change in UK Biobank (values > 0 indicatin proteins elevated in MS, values < 0 indicating proteins decreased in MS). Panel A shows comparison with the discovery cohort in Akesson *et al* 2023^12^. B: as per A, but for the replication cohort described in Akesson *et al* 2023. C: power curves showing the power to detect a difference between cases and controls at an alpha of 5% for a range of log-fold change values and cohort sizes. The dashed line indicates the log-fold change observed in UKB for neurofilament light chain, an established marker of MS. A range of scenarios is shown.

### Comparison with other autoimmune and neurodegenerative disorders

To determine whether any of the MS-associated protein biomarkers identified were specific to MS, we first explored the association between each of the 2911 plasma proteins and a range of autoimmune and neurodegenerative disorders. We reproduced several well-known associations (supplementary table 2), such as elevated beta-defensin in psoriasis^27^, interleukin-6 in ankylosing spondylitis^28^, TNF-alpha in rheumatoid arthritis^29^, IL-15 in SLE^30^, and neurofilament light chain in MND^31^, PD^5^, and AD^4^. The proteomic signature of MS showed no evidence of correlation with other autoimmune disease (supplementary figure 3). In contrast, we observed stronger correlations between autoimmune disorders with overlapping aetiological pathways, such as Crohn’s and UC (*P* < 0.001, correlation coefficient = 0.6).

Of the ten proteins found to be increased in MS, seven were strongly associated with another autoimmune disorder (*P*_Bonferroni_ < 0.05; MERTK, IL15, LILRB4, IL22, IL17RB, LAMP3, and CHGA, figure 3A-C). As expected, we found that elevation of NFL and GFAP was also associated with neurodegenerative disorders (figure 3A), with strong effects seen for Parkinson’s and Motor Neuron Disease. Most MS-associated biomarkers were not specific to MS: 46/49 (93.9%) showed at least nominal evidence of association (*P* < 0.1) in the same direction with another autoimmune disorder, and 34/49 (69.4%) were nominally associated with neurodegenerative disorder (figure 3B & C). The MS-associated decrease in Granzyme A (GZMA) was the only protein change apparently specific to MS (figure 3A), i.e. without any statistical evidence (i.e. *P* < 0.1) for a decrease in any other disorder tested (figure 3A & D). Reduction in plasma granzyme A was also observed in the external validation dataset (*P*_Discovery_ = 0.10, *P*_Replication_ = 0.02).

**Figure 3:**
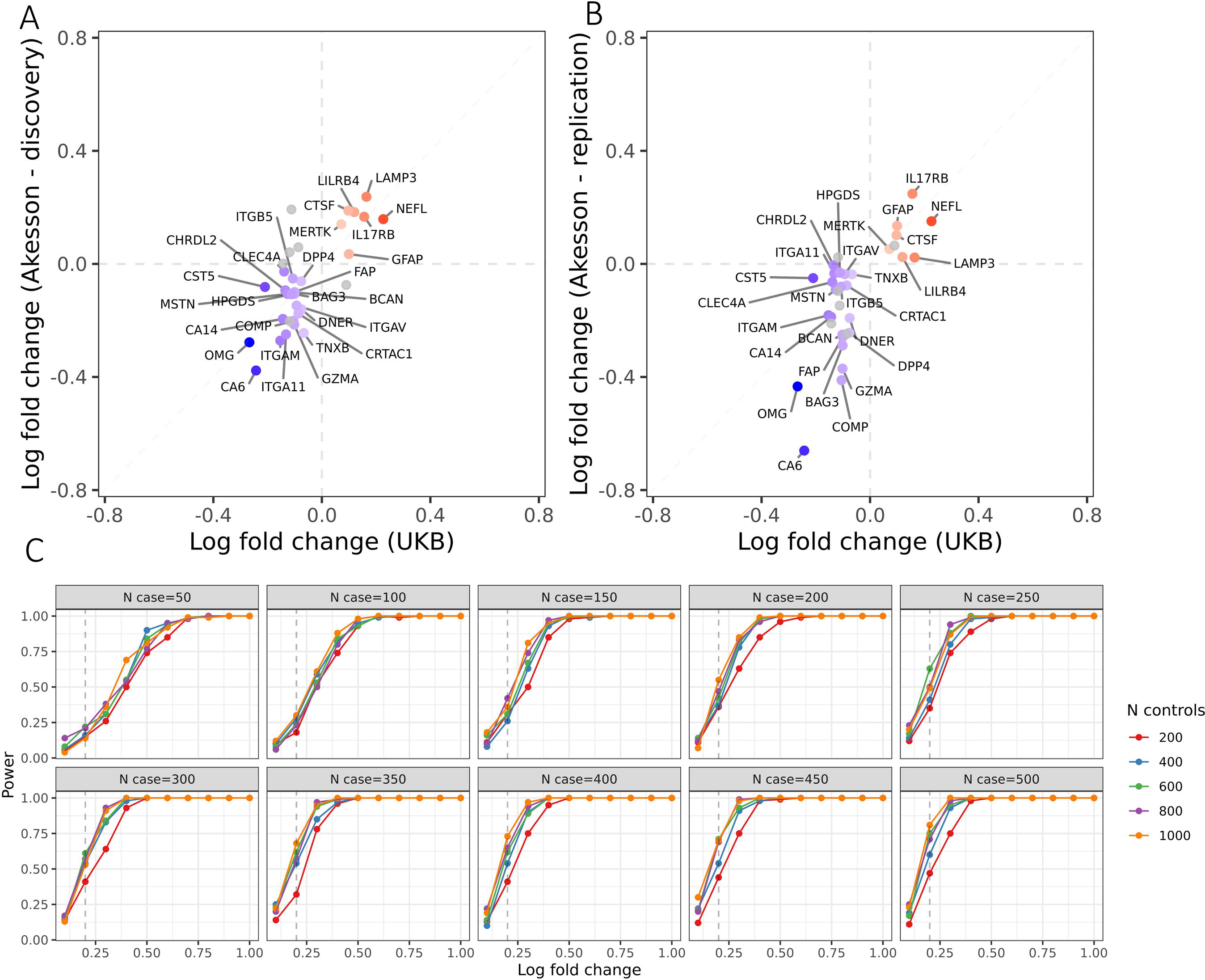
Decrease in plasma granzyme A may be specific to Multiple Sclerosis. A - forest plot showing the associations between the top proteins associated with MS (Bonferroni’s alpha < 0.001) and other autoimmune / neurological disorders. The x-axis shows the direction of effect (log-fold change) for the association of each protein with each disease outcome. The y-axis shows the protein tested. Each point is coloured by disease. MS-specific effects were defined as statistically-significant effects (Bonferroni’s alpha < 5%) in the MS cohort and did not show suggestive evidence (P<0.1) of the same direction of effect in any other disease tested. The only MS-specific effect according to this definition was the reduction in Granzyme A. B – venn diagram showing the overlap proteins found to be significantly upregulated (alpha < 0.05) in MS, autoimmune, and neurodegenerative (NDD) disorders. Note that in this figure overlap is defined as proteins achieving alpha < 0.05 for both/all conditions. C – as for panel B, but showing downregulated proteins. D – boxplots showing the normalised levels of plasma granzyme A across the disorders tested.

The reduction in plasma granzyme A could plausibly reflect the impact of MS treatment, rather than being a feature of the disease *per se*. To explore this possibility, we identified participants in UK Biobank who were receiving an MS disease-modifying therapy at the time of plasma sampling. The only MS disease-modifying therapy recorded for more than ten participants was interferon. We compared the proteomic profiles of MS patients taking interferon (n=29) versus those not taking interferon (n=378). Treatment with interferon was associated with upregulation of several known interferon-inducible proteins, including IFIT3, DDX58, and IFI30, but not with any alteration in granzyme A protein levels (supplementary figure 4, *P*=0.4).

### Proteomic correlates of brain atrophy and T2 lesion volume in MS

Next, we explored the relationship between the plasma proteome and radiological proxies for severity (T2 lesion volume and total brain volume) using MRI data available for a subset of the cohort. We first confirmed the expected increase in T2 lesion volume and decrease in brain volume in MS cases compared with healthy controls (N_MS_=113, N_Control_=3772, T2 lesion volume: median 10,417 mm^3^ vs 2885mm^3^, P < 2x10^-16^, figure 4A; Brain volume: median 1,465.1 cm^3^ vs 1,487.3 cm^3^, P < 0.003, figure 4B). To validate these measures, we demonstrated the expected negative correlation between T2 lesion volume and total brain volume (Spearman’s rho = -0.3, *P* = 0.0009, figure 4C), and the relationship between longer disease duration at the time of scanning and lower lower brain volume (rho -0.19, *P* =0.04, figure 4D). The impact of disease duration on T2 lesion load was not statistically significant (rho 0.08, *P* = 0.4). Male sex was associated with lower brain volume (*P* = 0.01) but did not have a statistically significant impact on T2 lesion volume (*P* = 0.2). Sensitivity analyses using a simplified model (i.e. adjusting for only age, sex, and proteomics batch) yielded similar results.

**Figure 4:**
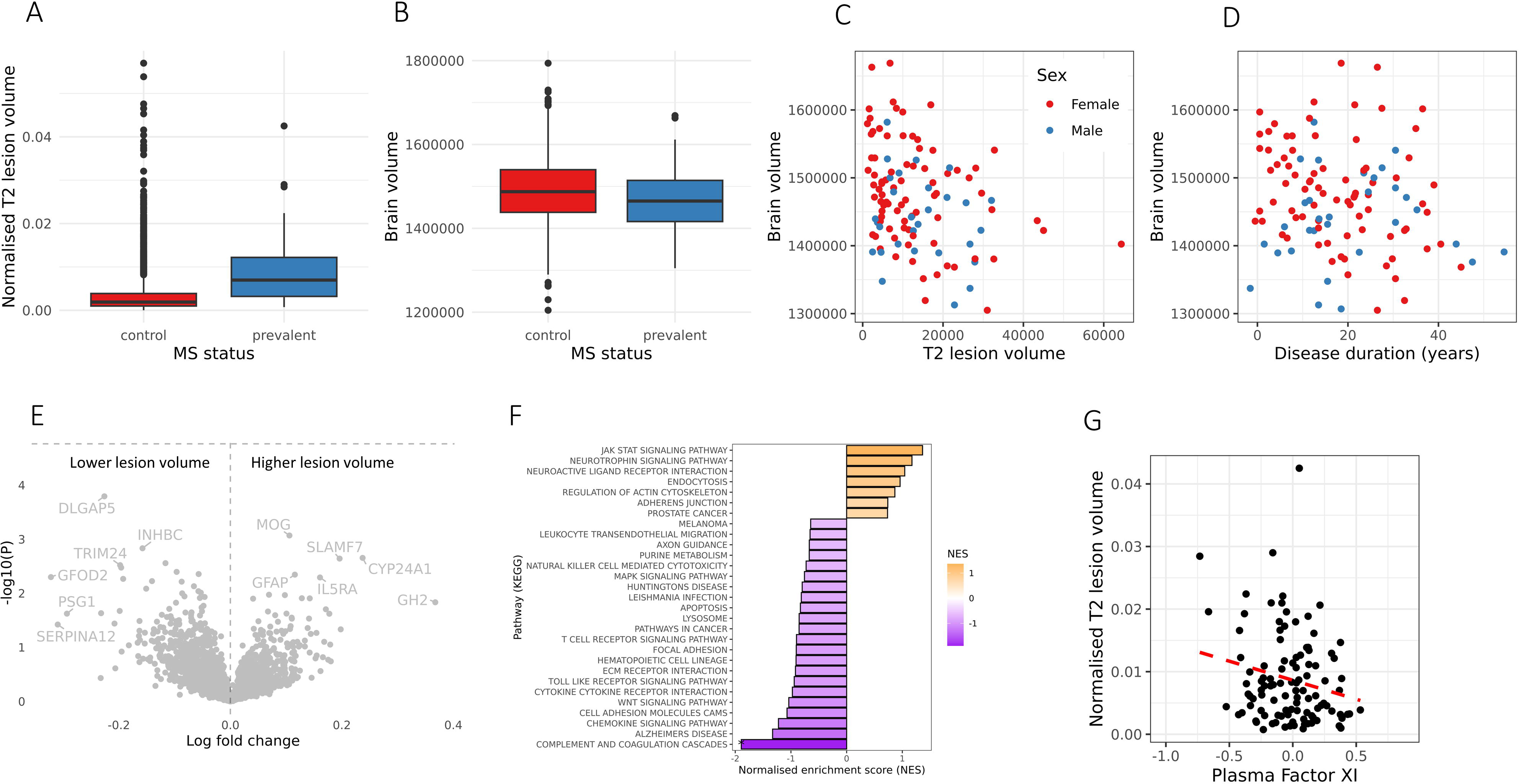
plasma proteomic correlates of MRI proxies for MS severity. A & B - boxplots showing the relationship between MS status and normalised T2 lesion volume load (A) or total brain volume (B) in the subset of MS cases and controls with MRI scans available. Normalised lesion load is shown as a proportion of total brain volume. Brain volume is shown in mm^3^. C & D – scatter plots showing the association of T2 lesion volume an brain volume in the MS cases (C) and between disease duration and brain volume (D), E - Volcano plots demonstrating the association between levels of plasma proteins and normalised T2 lesion volume in the MS cohort. F – GSEA results showing the enrichment of pathways associated with T2 lesion volume, highlighting the negative enrichment (i.e. inverse correlation) between plasma coagulation and clotting proteins and T2 lesion volume. G – scatter plot showing the association between plasma factor XI and normalised T2 lesion volume in the MS cases.

Although at an individual protein level there were no proteins associated with either T2 lesion volume or total brain volume at a stringent alpha of 5% (figure 4E, supplementary table 3), we found suggestive evidence (*P* < 0.05) for several plausible associations. For example, for T2 lesion volume, we found suggestive positive associations with the established markers of neuroaxonal damage NFL and GFAP, the myelin protein MOG, proteins involved in modulating the immune response (SLAMF7, IL5RA), and the vitamin D-metabolising enzyme CYP24A1. At a pathway level, we observed strong evidence (FDR < 5%) for a role of proteins involved in complement and coagulation (figure 4F), with negative enrichment seen for T2 lesion volume and positive enrichment for brain volume (i.e. higher levels of these proteins appear to be ‘protective’). This effect was driven by alterations in 21 proteins (supplementary table 3), of which only one, factor XI, achieved suggestive evidence (*P* < 0.05) with both T2 lesion volume and brain volume, with higher plasma factor XI associated with higher brain volume (*P* = 0.008) and lower T2 lesion volume (*P* = 0.03, figure 4G). Plasma factor XI was not associated with T2 lesion volume (*P =* 0.3) or brain volume (*P* = 0.8) in the healthy controls (N = 3644), suggesting that this association is unlikely to reflect nonspecific brain health (supplementary table 3).

## Discussion

Using data from ∼50,000 UK Biobank participants, we describe alterations in the plasma proteome of patients with Multiple Sclerosis. We report that people with MS have higher plasma levels of neuronal damage markers (NFL, GFAP), cytokines/cytokine receptors (IL15, IL17RB, IL22), and lower levels of several proteins, including integrins and cystatin D, compared with healthy controls. We find that most of these associations show evidence of replication in an external dataset. While most of these alterations are also seen in other autoimmune or neurodegenerative disorders, we find that the decrease in plasma granzyme A, a serine protease released by NK cells as part of the degranulation response, is relatively specific to MS, and is not explained by treatment with interferon. Finally, using MRI scans from a subset of the cohort, we report that T2 hyperintense lesion burden shows suggestive associations with plasma NFL, MOG, and SLAMF7, and that higher levels of plasma proteins involved in coagulation, such as factor XI, are associated with preserved brain volume and lower T2 lesion load.

This is the first study to evaluate MS-associated changes in the plasma proteome using biobank-scale data. The key strengths of our study are, firstly, the very large number of healthy controls, minimising the risk of false positive findings, and the broad coverage of the proteome afforded by the Olink platform. Plasma NFL elevation in MS is a well-established biomarker of neuroaxonal damage, which can be detected early in the course of MS, and may be a useful biomarker for tracking disease activity and progression over time^2^. The fact that plasma NFL was the protein most strongly and consistently associated with MS in our study across a range of sensitivity analyses is a reassuring positive control^16^.

In addition to the established markers of neuroaxonal damage NFL and GFAP, we identify elevation of several plasma proteins in MS patients, including cytokines and cytokine receptors (IL22, IL17RB, and IL15), proteins involved in lysosomal processing (LAMP3, CTSF), and regulators of monocyte/microglial function (MERTK, LILRB4)^12,32–36^. While these findings suggest a variety of plausible targets for therapeutic intervention in MS, most of these alterations were observed in other autoimmune disorders and/or neurodegenerative conditions, suggesting that they are unlikely to be of significant diagnostic value in distinguishing MS from alternative conditions. The reduction in granzyme A was apparently specific to the MS cohort, and was observed independently in an external dataset^12^. Granzyme A is a serine protease released by degranulating cytotoxic cells (such as CD8+ T cells and NK cells)^37^. These data are somewhat paradoxical in the context of previous findings suggesting patients have higher levels of granzyme A in the CSF during relapse^38^, and describing higher proportions of age-associated GZMA+ T cells than controls^39^. Further work is required to clarify the relationship between membrane-bound granzyme A, plasma granzyme A, and CSF granzyme A. Plausible interpretations of these data are that this may be an effect of treatment – which we attempt to address – and potentially a feature of accelerated immune ageing in MS.

Despite the low statistical power to detect associations between plasma proteins and radiological proxies of MS severity (T2 lesion burden and brain volume), we detect suggestive associations with high plausibility. For instance, we show nominal (*P* < 0.05) associations between T2 lesion volume and plasma NFL, MOG, and surface antigen CD319 (SLAMF7), an MS susceptibility gene^40^ which also showed suggestive association with brain volume. The positive control of NFL reinforces the validity of this approach and suggests that novel targets could be discovered with an increase in sample size. At a pathway level, we observe evidence implicating the complement and clotting pathways as ‘protective’ against higher lesion load and brain volume loss. Although there are several studies reporting clotting abnormalities in MS patients^41^, most observational studies have reported the opposite direction of effect (i.e. higher plasma levels of clotting factors correlating with inflammatory disease activity^42^), and inhibition of factor XI – the most suggestively associated protein in our study – was beneficial in a rodent model of MS^43^. Replication in well-phenotyped longitudinal cohorts is required to explore this intriguing effect.

There are some important limitations to this study. First, we use data from a single cohort assayed with a single technology (Olink), and so although we compare our findings with published data, we are unable to test our results in a genuine replication cohort. Second, the major drawback of using UK Biobank to address this question is that this is an all-purpose dataset not specifically designed for MS. We are therefore unable to explore MS phenotypes in granular detail - for instance, we are unable to explore the relationships between the proteome and EDSS, relapse rate, treatment responsiveness, or more MS-specific radiological outcomes such as new/expanding lesion load. There is also some risk of misclassification of cases and controls inherent in using electronic healthcare records and self-reported diagnoses rather than clinically-definite MS diagnosis, although we have previously shown that this cohort is demographically similar to clinical MS cohorts^17^. While we attempt to control for treatment, a small proportion of cases have any disease-modifying therapy recorded, and we are unable to distinguish untreated patients from those with missing data. Many of our findings could be explained by the effects of treatment, such as the reduction in Granzyme A. Third, as this study is cross-sectional and includes participants with established MS, it is challenging to deconvolve the impact of treatment, disease duration, disease stage at the time of sampling, and other time-varying parameters. We mitigate this to some extent by accounting for a variety of important confounders, such as age, sex, BMI, and deprivation status, but the plasma proteome is likely highly dynamic, and a cross-sectional snapshot is a gross simplification of this complexity. Fourth, it is now established that participants in UKB are a highly selected population, and so it is plausible that the MS cohort we analysed here represents an unusually ‘healthy’ selection of MS patients^44^. There is an even greater degree of selection bias determining which participants enrol for the imaging substudy, and so the risk of collider bias is even greater in the analysis of proteomic correlations of MRI outcomes. This phenomenon could explain some of the apparently paradoxical results we observed, such as the reduction in Granzyme A in MS patients and the protective effect of increased plasma clotting factors. Finally, while the Olink assays give excellent coverage across the proteome, it still only measures a finite set of proteins and may miss a variety of interesting proteins either not present on the panel or which have undergone post-translational modifications, in contrast to the relatively unbiased nature of mass spectrometry.

In summary, we perform the largest analysis of the plasma proteome in MS, replicating known biomarkers for diseases such as NFL and GFAP, suggesting plausible targets for therapeutic intervention, discovering a new disease-specific negative biomarker (GZMA), and suggesting an association between plasma coagulation factors and MRI outcomes in MS. While these findings require external replication, they demonstrate the power of biobank-scale datasets for discovering how the plasma proteome is altered in Multiple Sclerosis. Ultimately, this avenue of research could yield new drug targets, new insights into disease biology, and provide an adjunct to existing methods for individual-level prognosis in MS.

## Acknowledgements

BMJ is funded by a Medical Research Council (MRC) Clinical Research Training Fellowship (CRTF) jointly supported by the UK MS Society (BMJ; grant reference MR/V028766/1), and by Barts Charity. The Preventive Neurology Unit is supported by a grant from Barts Charity.

## Author contributions

BJ conceived of the study, conducted the analysis, and wrote the first draft. SW helped with curation and cleaning of the dataset. PP, NV, GG, SW, and RD critically reviewed the manuscript for important intellectual content. All authors reviewed and approved the final draft.

## Competing interests

BMJ, NV, PP and SW report no relevant competing interests.

GG reports personal compensation for participating in advisory boards in relation to clinical trial design, trial steering committees, and data and safety monitoring committees from: Abbvie, Atara Bio, Biogen, Canbex, Sanofi-Genzyme, Genentech, GSK, MSD, Merck-Serono, Novartis, Roche, Synthon BV and Teva. RD has received research support from Multiple Sclerosis Society UK, Horne Family Foundation, Barts Charity, Merck, Biogen and Celgene; consultancy fees from Roche, Novartis, Janssen and Biogen (all payments made are institutional and used to support research/educational activities); honoraria for lectures, speaking etc. from Biogen, Roche, Sanofi-Genzyme, Merck, Novartis, and Janssen; support for attending meetings and/or travel from Novartis, Biogen, Roche and Janssen (all payments made are institutional and used to support research/educational activities).

## Supplementary material

Supplementary material is available is enclosed.

## Notes

### Competing Interest Statement

The authors have declared no competing interest.

### Author Declarations

UK Biobank has approval from the North West Multi-centre Research Ethics Committee (MREC) as a Research Tissue Bank (RTB) approval. This approval means that researchers do not require separate ethical clearance and can operate under the RTB approval (there are certain exceptions to this which are set out in the Access Procedures, such as re-contact applications).

### Summary of Updates

We have included more proteomics data from UK Biobank, corrected an issue with the covariate data, performed additional sensitivity analyses and power calculations, and included some discussion of the impact of treatment.

