## Supplementary figures for "Plasma proteomic profiles of UK Biobank participants with Multiple Sclerosis"

**Supplementary materials for “Plasma proteomic profiles of UK Biobank participants with Multiple Sclerosis”**

Supplementary figure 1: disease counts for prevalent and incident occurrences of neurodegenerative disorders, autoimmune disorders, and MS in the UK Biobank proteomics cohort. Cases are coloured according to whether they were reported more than two years after recruitment (incident), or before that point (prevalent). As expected given that recruitment took place between the ages of 40 and 69, most MS cases were identified prior to recruitment, whereas most neurodegenerative diagnoses have been reported during follow-up.

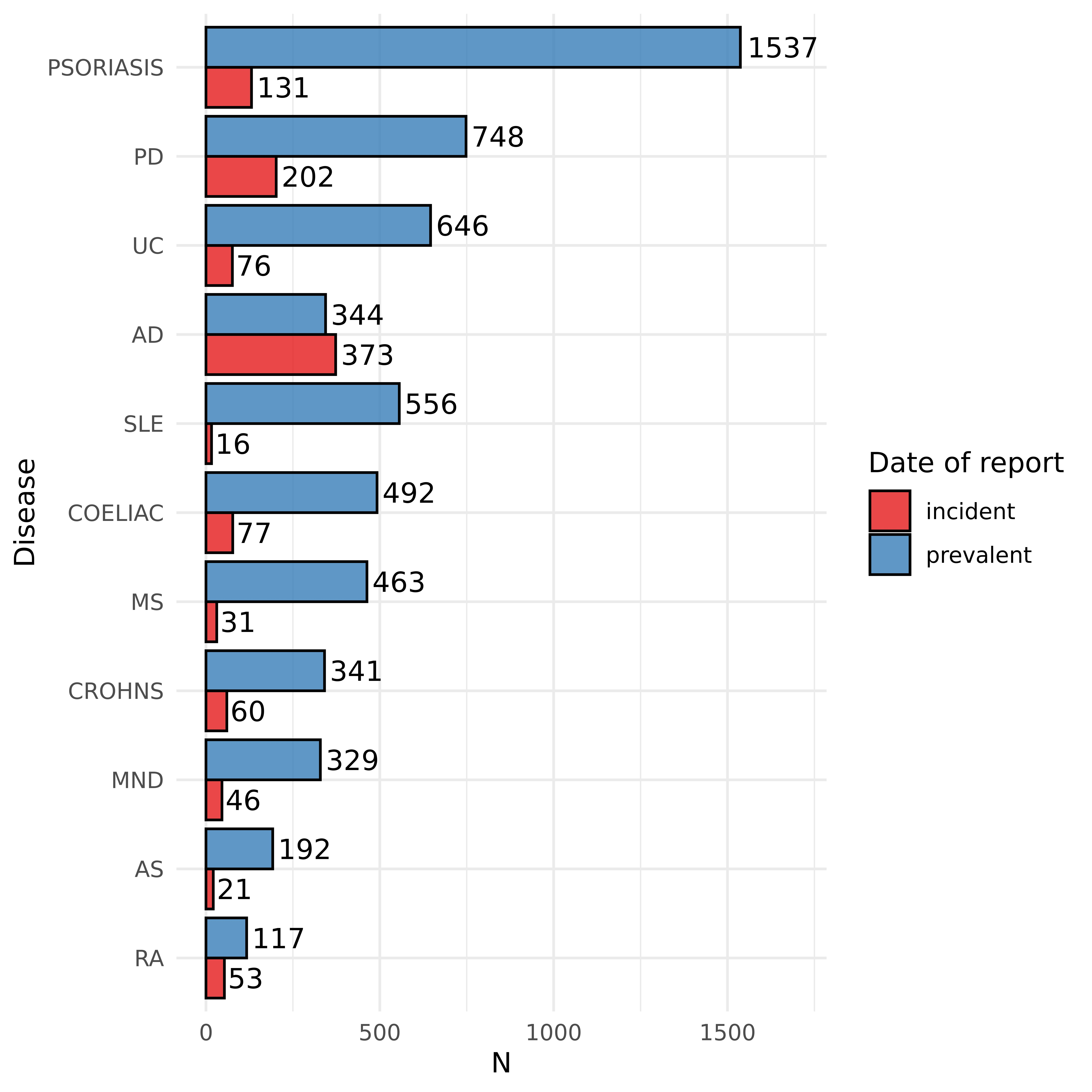

Supplementary figure 2: individual filtering and quality control. From an initial cohort of 52,705 UKB participants with proteomic data, 380 prevalent MS cases and 38,263 controls were included in the primary analysis. A subset with MRI data - 105 prevalent MS cases - were included in the analysis of MRI lesion volume.

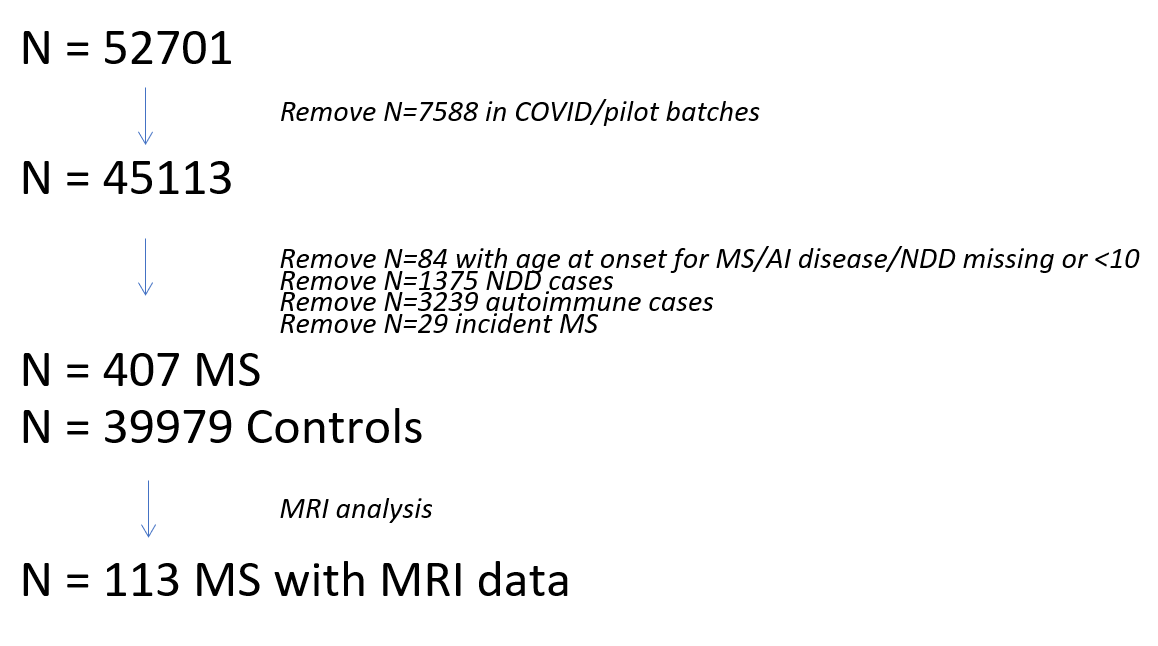

Supplementary figure 3: correlation plot showing the correlation between protein-disease associations (beta coefficients) for the range of disorders tested. Cells are coloured by the strength of the correlation coefficient, ranging from a strong negative correlation (orange) to a strong positive correlation (blue). Asterisks indicate the statistical significance of the correlation. * P < 0.05, ** P < 0.01, *** P < 0.001.

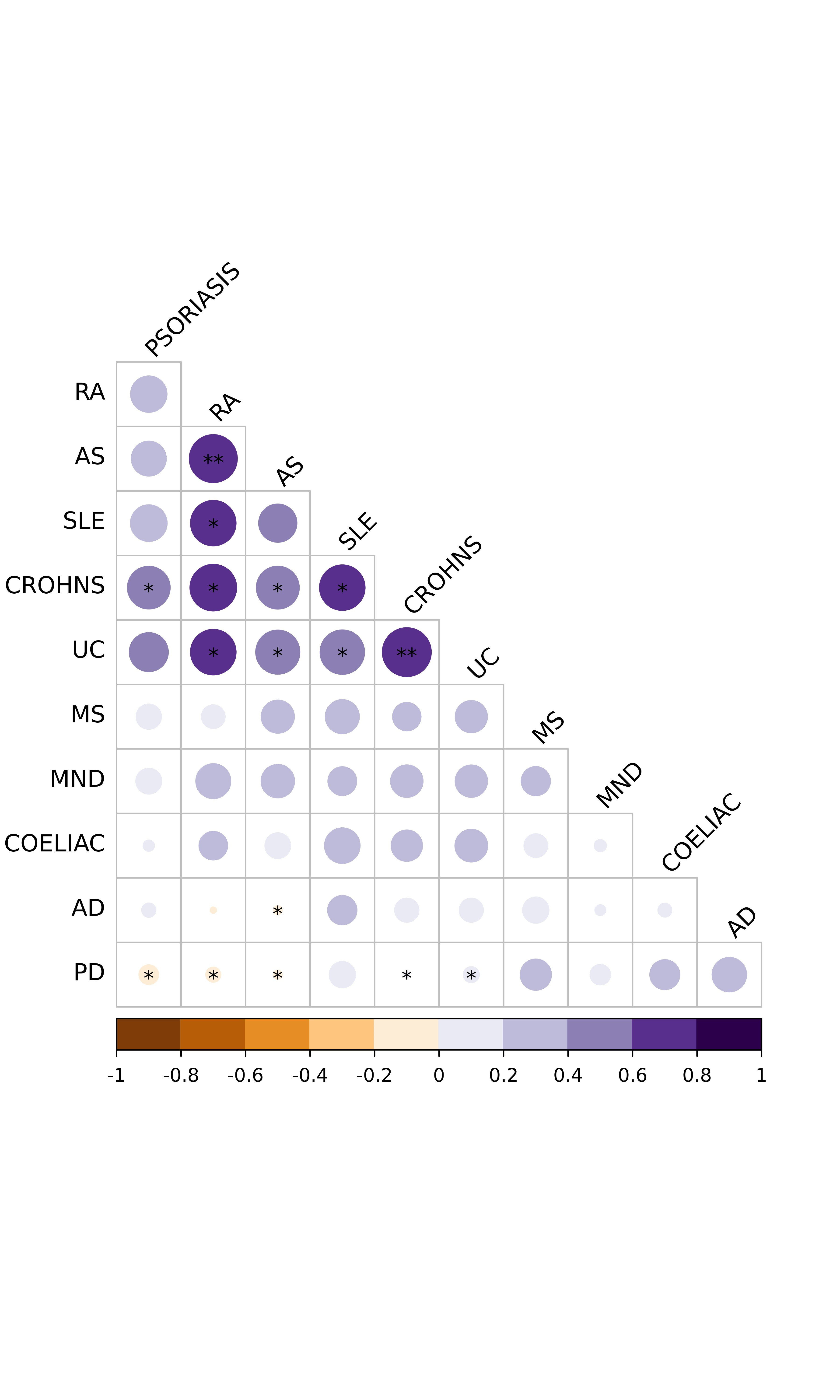

Supplementary figure 4: Volcano plot showing the association of plasma protein levels with current interferon treatment in the prevalent MS cohort (n=29 on treatment vs. 378 controls). Proteins coloured in red are upregulated in those on interferon, those in blue downregulated, and those in grey not statistically significant at an alpha of 0.05.

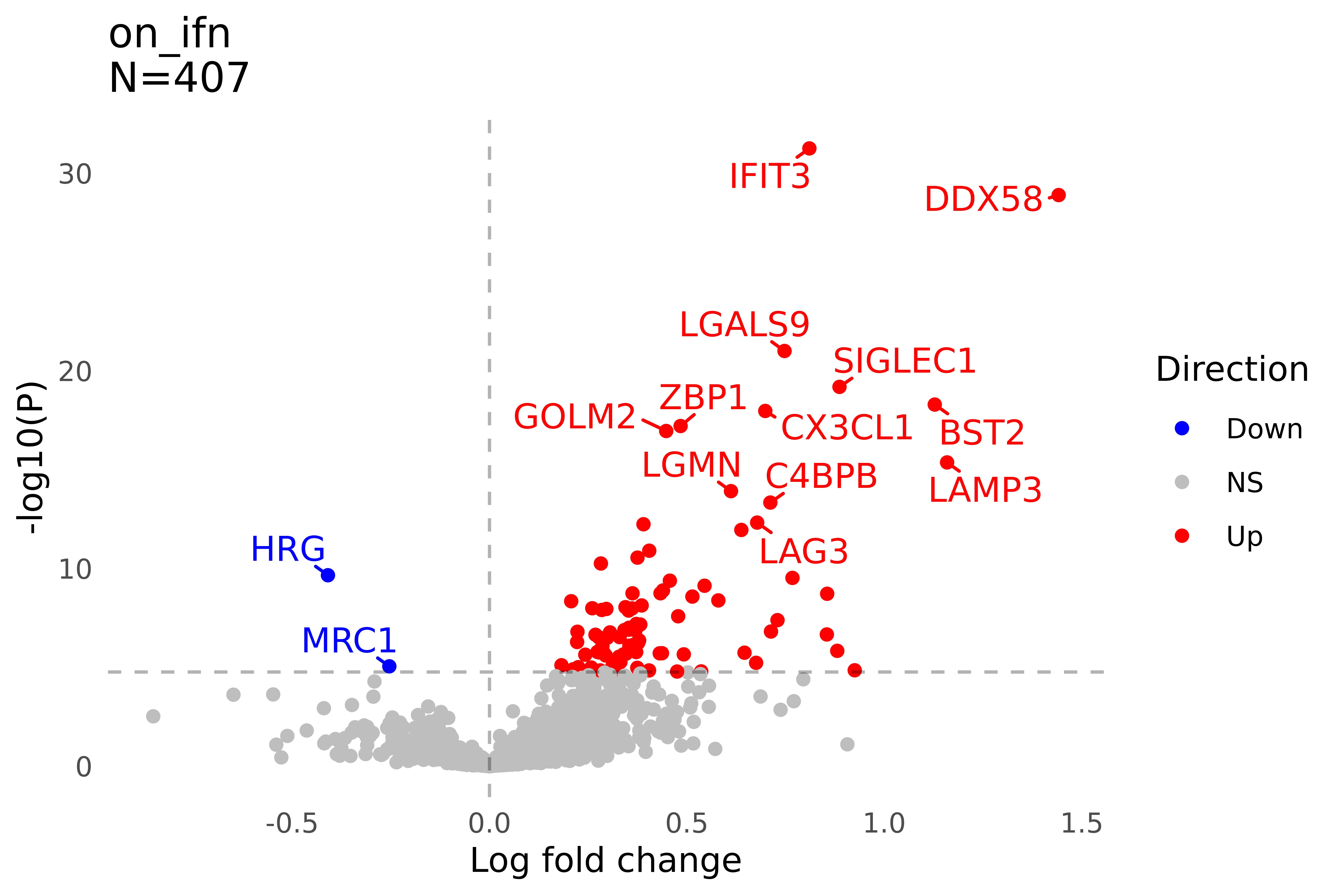
